# TO DETERMINE THE KNOWLEDGE OF MIDWIVES ON UTILIZATION OF PARTOGRAPH AT SELECTED HOSPITALS IN KENYA

**DOI:** 10.1101/2023.09.06.23294812

**Authors:** Mary wangeci kanyottu, Jane karonjo, Anna karani

**Affiliations:** Mount Kenya University; University of Nairobi; mount kenya university

## Abstract

Partograph is graphical tool presentation of key data during the progress of labor. The observations of the mother are interpreted by a midwife or an obstetrician to make conclusions to whether the condition of the mother and fetus is fine. This tool alerts the skilled birth attendants of pending risk of the mother or fetus through the observations and action taken without delay. Worldwide the Maternal Mortality Rate is 400/ 100,000 live births. A woman from developing world is 100 times more probable to die during pregnancy or child birth than a woman from a developed country. The purpose of the study is to develop a model to promote partograph utilization by midwives during management of mothers in labour in selected hospitals in Kenya so as to reduce Maternal Mortality rate related to delivery.

## 1.0 Introduction

### 1.1 Background of the Study

Partograph is graphical tool presentation of key data during the progress of labor. It is a vital tool to assist midwives to detect whether labor is normal or abnormal and to warn the woman with the help of the midwife immediately the signs of fetal distress. Bedwell et al. (2017) noted that maternal and fetal complications due to prolonged labour were less common with labour progress observed by the midwives using a partograph. Partograph is used by a trained skilled birth attendant to record the observations to determine mother’s condition as well as that of the fetus, and the progress of labour in totality. The midwives should be able to make interpretation of the observations and give feedback to the mother of the progress of labour. In this regard, it is advisable to use the tool while taking care of mother in labour, be it at the facility or at home (WHO, 2018).

The partograph is a chart that helps to find out normal or abnormal labour progress that is prolonged or obstructed. Partograph may alert midwife to any foetal distress. Its main purpose is providing a pictorial labours summary that will warn the birth attendant to abnormalities in labour progress and maternal or fetal safety (Lavender, 2013). According to (WHO 2014). Partograph has three distinctive parts where observations allied to labour progress, maternal and foetal condition are recorded. The Partograph is clearly demarcated to of labour complications for example poor labor progress, foetal distress, prolonged or obstructed labour (WHO, 2018).

Partograph provides health care givers labor’s pictorial overview to ensure that there is early identification as well as diagnosis of the complications associated with labour. WHO recommends partograph in monitoring labour and delivering, with the goal of improving health care and reduce maternal in addition to fetal morbidity or death (WHO, 2015). According to WHO assessment of outcome of the use of partograph, it advocated skilled midwives to use it as an essential tool in the managing labour. WHO proposes usage of partograph for monitoring maternal labour globally (WHO, 2018). The main objective is to prevent complications associated with labour so as to lower the rates of maternal and perinatal morbidity and mortality.

There are major reasons why skilled birth attendants should monitor a mother in labour so carefully so as to detect complications early and act urgently. Berhan (2014) argues that a well progressing labour requires less help of a health professionals compared to labour identified to be progressing abnormally. Recording partograph results in labour allows the attendant to be informed labour is abnormal, and if a mother should be referred to the closest higher level facility for more medical or intervention.

World Health Organization still describes partograph as a graphical tool monitoring labor progress where the important details of the mother as well as fetus are documented (WHO, 2017). The tool currently is still being recommended by WHO to be well utilized. The tool was originally introduced as a system of early warning to detect abnormal labour as detected by skilled birth attendants taking care of women in labour. Many cases of maternal complications and deaths are due of obstructed as well as prolonged labour which can be prevented through an affordable and various cost-effective health interventions critical tool like the use of partograph (Lavender et al, 2014). When proper monitoring of labour by use of partograph is embraced, then maternal mortality will drastically be reduced (Asibong et al., 2014). The partograph indicates when augmentation is required, and indicate possibility of cephalopelvic disproportion just prior to labour obstruction. Partograph increases the quality as well as consistency while observing the mother and the new born.

The tool that WHO recommends has worked in several of places globally (WHO, 2017). Partograph has been more operational in averting prolonged labour and improving neonatal outcome. The WHO partograph starts only in active labour stage, with 4 cm of cervix or more dilated. Nevertheless, it is a tool whose effectiveness is dependent on healthcare professional using it.

The midwife together with other skilled birth attendants taking care of the mother in labour should be able to chart the observations that they take. Maternal pulse rates should be taken like after every half an hour, blood pressure and temperature four (4¬) hourly, urine production and dipstick testing for protein, ketones (if accessible) just as glucose subsequent to voiding. All liquids and medications regulated are similarly checked and recorded. On the off chance that the discoveries become irregular, expanded recurrence of perception and testing was required, and intercession may be executed. The woman in labour should be taken care of well, monitored carefully to ensure she is safe with no labour complications (Hodnett et al 2012).

For fetal wellbeing, the fetal heart rate checked for 1 minute every 15–30 minutes after a contraction during the first stage and recorded. In secondary stage every 5 minutes fetal heart rate should be taken and noted down on the tool. During the second stage, if abnormalities are recorded, urgent delivery should be considered by encouraging the mother to push.

The Liquor status should be observed whether clear, meconium recolored (thick or slim), grisly or missing. Thick meconium recommends fetal misery, and closer observing of the embryo is demonstrated. This ought to be checked after like clockwork. The recurrence, length and quality of uterine compressions (surveyed by palpation) ought to be noted after at regular intervals. At long last the vaginal assessment ought to be done at regular intervals to evaluate cervical dilatation, plummet of the fetal head, and trim of skull bones. Progressively visit assessment is possibly attempted whenever demonstrated in order to confirm cervical dilatation.

### 1.2 Statement of the Problem

Mothers who become pregnant and attend Ante Natal Clinic (ANC) are prepared well for delivery under the skilled birth attendants‟ care. The mothers end up with successful labour outcome with minimal delivery complications if they attended recommended minimum of eight ANC visits by World Health Organization (WHO, 2017). In countries where mothers don’t attend ANC have high maternal mortality rates. This is seen is a global burden, with at least 585 000 women dying annually due to pregnancy related complications and childbirth (Kitila et al. 2014). If skilled birth attendants are able to detect maternal and foetal complications early through the use of partograph as tool to monitor labour, the death associated to child birth should be minimized (Bazirete et al, 2017). The UN report indicate that by the year 2030 the global MMR is expected to be reduced to < 70/100 000 live births and neonatal mortality by 12/1000 live births to attain the Sustainable Development Goals by 2030 (Alkema et al. 2016).

## 2.0 Literature Review

### 2.1 Midwives’ Competencies on Utilization of Partograph

Competency goes deeper beyond mere knowledge, skills or attitudes when the midwives get experience in managing mothers in labour. It includes the ability of behaviours, as well as characteristics that a person needs to perform a duty, or occupational roles successful (Sturgess, 2012). Skill is an ability besides capacity obtained by deliberate as well as sustained effort to adaptively and smoothly perform complex activities involving things technical skills, cognitive skills, and/or interpersonal skills so as to counter complex demands, by drawing on and mobilizing psychosocial resources in a given context. Competency may likewise be described as a measurable pattern of skills, knowledge, attitudes reached when the learner is capable of performing what was taught and demonstrated and keeps practicing till the level of proficiency.

Midwives as part of skilled birth attendants are expected to utilize partograph as a labour monitoring tool and be able to chart and interpret the observations whether on paper or digital. Action should be taken immediately the midwife realizes that the observations are not normal. An Obstetrician on call should be informed if not in labour ward to review the condition of the mother and foetus and take appropriate action (Lavender et al, 2013). If the mother is not in a tertiary level facility with a caesarian section (CS) can be done urgently, the referral should be done without delay. The following should be plotted on the partograh by the skilled birth attendant:

### 2.2 Recording the Condition of the Fetus Assessment and labour management

After every examination an assessment must be conducted and documented on the partograph. The findings of observations recorded on the partograph was interpreted keenly as part of monitoring of labour progress.

Recording the time on the partograph

The partograph will also have a record of time, to the nearest half hour, each time an observation is documented, medication is provided, an assessment is made or any prior management is changed (Myles Textbook for Midwives, 2014).

## 3.0 Research Methodology and Study Design

The research will employ a descriptive cross-sectional study design was conducted among the midwives working in labour wards from the selected hospitals. Data was collected using both quantitative and qualitative methods including, questionnaire and key informant, Focus Group Discussion (FGD) interview was administered to various labour ward midwives.

Participants who was randomly selected from the four selected County referral hospitals. The study will use questionnaires for working midwives and Focus Group Discussion of selected midwives working in labour ward and supervisors to collect data. The questionnaires will have both open ended and closed ended type of questions which was developed and administered to collect data from the respondents.

Data collected was assessed for accuracy then was cleaned, edited, re-classified and coded before entry into Statistical Package for Social Services (SPSS) version 24 for analysis.

Means & standard deviations was used to analyzed the quantitative continuous variables like age, marital status, education level, knowledge, religion on partograph. Chi square was used to determine associations between level of education and the danger signs of complicated labour, plan of intervention of danger signs, obstetric emergencies. Midwives t tests was used to analyze associations between age, level of education, experience duration worked in labour ward.

## 4.0 Results and finding

### 4.1 Descriptive statistics on the knowledge of midwives on utilization of partograph at selected hospitals in Kenya

Table 1 below indicates the descriptive analysis on the knowledge of midwives on utilization of partograph at selected hospitals in Kenya.

**Table 1:** Knowledge of midwives on utilization of partograph at selected hospitals in Kenya.

|  | Yes |  | No |  |
| --- | --- | --- | --- | --- |
|  | F | % | F | % |
| Have you ever used a Partograph before? | 129 | 98.5% | 2 | 1.5% |
| A chart for monitoring of labour by doctors | 29 | 22.1% | 102 | 77.9% |
| A chart for monitoring of labour by midwives | 82 | 62.6% | 49 | 37.4% |
| A complex tool with pictorial overview of labour for the use by midwives | 41 | 31.3% | 90 | 68.7% |
| A simple graphic tool to record progress of labour and salient conditions of mother and fetus against time in hours | 117 | 89.3% | 14 | 10.7% |
| Did you receive any training on the use of partograph? | 128 | 97.7% | 3 | 2.3% |
| If yes, where have been trained on the use of partograph? from Colleague | 64 | 48.9% | 67 | 51.1% |
| If yes, where have been trained on the use of partograph? from medical doctor | 3 | 2.3% | 128 | 97.7% |
| If yes, where have been trained on the use of partograph? from school | 92 | 70.2% | 39 | 29.8% |
| If yes, where have been trained on the use of partograph? in service training | 82 | 62.6% | 49 | 37.4% |
| Are you willing to have in service training on the use of Partograph? | 130 | 99.2% | 1 | 0.8% |
| The Partograph is one of the tools for implementing safe motherhood | 100 | 76.3% | 31 | 23.7% |
| The partograph will reduce maternal deaths | 131 | 100.0% | 0 | 0.0% |
| The partograph will reduce new born deaths | 131 | 100.0% | 0 | 0.0% |
| In a normal progress of labour, the graph/plot on the Partograph should fall on the alert line | 106 | 80.9% | 25 | 19.1% |
| In a normal progress of labour, the graph/plot on Partograph should fall on the left of alert line | 73 | 55.7% | 58 | 44.3% |
| In a normal progress of labour, the graph/plot on Partograph should fall on the right of alert line | 49 | 37.4% | 82 | 62.6% |
| In normal labour, a woman has got 3 contractions in 12 hours | 11 | 8.4% | 120 | 91.6% |
| Indicates appropriate action must be taken | 121 | 92.4% | 10 | 7.6% |
| Allows time for the woman to be adequately assessed for appropriate intervention | 117 | 89.3% | 14 | 10.7% |
| Continuous observation till delivery | 92 | 70.2% | 39 | 29.8% |
| Upon diagnosis of labour | 77 | 58.8% | 54 | 41.2% |
| While the woman is still in labour | 124 | 94.7% | 7 | 5.3% |
| After delivery of the baby | 56 | 42.7% | 75 | 57.3% |

#### Source Field Data (2022)

According to table 5 above, most of the respondent have used partograph before with a rate of 129(98.5%) while only 2(1.5%) that had not used partograph before.

Most defined partograph as a simple tool with pictorial overview progress of labour and salient conditions of mother and fetus against time in hours with a rate of 117(89.3%) while 14(10.7%)did agree with these definition while those defined partograph by midwives were 82(62.6%) and those who did not with these definition were 49(37.4%), those who defined partograph as a complex tool with pictorial overview of labour for the use by midwives were 41(31.3%) and those who did not agree with these definition were 90(68.7%) and finally those who defined partograph as a chart for monitoring of labour by doctors were the lowest with a rate of 29(22.1%) and those who did not agree with these definition were 102(77.9%).

Many are those who received training on the use of partograph with a rate of 128(97.7%) while those who did not receive training were only 3(2.3%).

Training was mostly from school with a rate of 92(70.2%) while 39(29.8%) did not receive from school, 82(62.6%) received from in-service training while 49(37.4%) did not receive from in service training, 64(48.9%) received training from colleagues while 67(51.1%) did not receive from colleague and lastly 3(2.3%) received from medical doctor while 128(97.7%) did not receive from medical doctor.

Those respondents that were willing to receive in service training on the use of partograph were 130(99.2%) and only 1(0.8%) was not willing to receive in service training.

Every respondent has the understanding about the partograph as partograph will reduce maternal deaths and reduce new born deaths as the respondent rate was 131(100%), those who had understanding as partograph is one of the tools for implementing safe motherhood were 100(76.3%) while those who did have these understanding were 31(12.7%), 106(80.9%) had the understanding about the partgraph as in a normal progress of labour, the graph/plot on the partograph should fall on the alert while 25(19.1%) did not have these understanding, 73(55.7%) had understanding as in a normal progress of labour, the graph/plot on partograph should fall on the left of alert line while 58(44.3%) did not have these understanding, 49(37.4%) had the understanding as in a normal progress of labour, graph/plot on partograph should fall on the right of alert line while 82(62.6%) did not have these understanding and finally 11(8.4%)had the understanding as in normal labour, a woman has got 3 contractions in 12 hours and 120(91.6%) did not have these understanding.

Respondent who agreed with the function of the action line on the partograph as an indicator of appropriate action must be taken were 121(92.4%) and 10(7.6%) did not agree with these function, 117(89.3%) respondent agreed that it allows time for the woman to be adequately assessed for appropriate intervention while 14(10.7%) did not agree with these function and 92(70.2%) agreed that is a continuous observation till delivery while 39(29.8%) did not agree with these function.

Mostly in the hospital/practice information entered onto the partograph while the woman is still in labour with a rate of 124(94.7%) while those who did not enter during these time were only 7(5.3%), those who entered upon diagnosis were 77(58.8%) while those who did not enter during these time were 54(41.2%) and finally those entered information onto the partgraph after delivery of the baby were 56(42.7%) while those who did not enter during these time were 75(57.3%).

In contrast, Yisma et al. (2013) found that 60.0% of respondents agreed that, in normal labor progress, the plot on the partograph falls to the left of the alert line, while 25.1% disagreed and 14.9% did not know. Among 195 respondents, 66.2% agreed that it would fall on the alert line, while 17.9% disagreed and 15.9% did not know. Additionally, 34.4% of respondents agreed that, in normal labor progress, the plot on the partograph would fall to the right of the alert line, while 49.2% disagreed and 16.4% did not know. Overall, 52.3% of respondents had fair knowledge of the partograph, while 39.0% had good knowledge, and 8.7% had poor knowledge. Furthermore, only 25% of public health officers had a good level of knowledge, while 53.3% of midwives had a good level of knowledge, and the majority of nurses (68.4%) were rated as having a fair level of knowledge about the partograph.

### 4.2 Inferential statistics on the knowledge of midwives on utilization of partograph at selected hospitals in Kenya

**Table 2:** One Sample analysis on the knowledge of midwives on utilization of partograph at selected hospitals in Kenya.

| **One-Sample Statistics** |  |  |  |  |
| --- | --- | --- | --- | --- |
|  | N | Mean | Std. Deviation | Std. Error Mean |
| Have you ever used a Partograph before? | 131 | 1.05 | .369 | .032 |
| A chart for monitoring of labour by doctors | 131 | 1.78 | .417 | .036 |
| A chart for monitoring of labour by midwives | 131 | 1.37 | .486 | .042 |
| A complex tool with pictorial overview of labour for the use by midwives | 131 | 1.69 | .465 | .041 |
| A simple graphic tool to record progress of labour and salient conditions of mother and fetus against time in hours | 131 | 1.11 | .310 | .027 |
| Did you receive any training on the use of partograph? | 131 | 1.02 | .150 | .013 |
| If yes, where have been trained on the use of partograph? from Colleague | 131 | 1.51 | .502 | .044 |
| If yes, where have been trained on the use of partograph? from medical doctor | 131 | 1.98 | .150 | .013 |
| If yes, where have been trained on the use of partograph? from school | 131 | 1.30 | .459 | .040 |
| If yes, where have been trained on the use of partograph? in service training | 131 | 1.37 | .486 | .042 |
| Are you willing to have in service training on the use of Partograph? | 131 | 1.01 | .087 | .008 |
| The Partograph is one of the tools for implementing safe motherhood | 131 | 1.24 | .427 | .037 |
| The partograph will reduce maternal deaths | 131 | 1.00 | .000<sup>a</sup> | .000 |
| The partograph will reduce new born deaths | 131 | 1.00 | .000<sup>a</sup> | .000 |
| In a normal progress of labour, the graph/plot on the Partograph should fall on the alert line | 131 | 1.19 | .394 | .034 |
| In a normal progress of labour, the graph/plot on Partograph should fall on the left of alert line | 131 | 1.44 | .499 | .044 |
| In a normal progress of labour, the graph/plot on Partograph should fall on the right of alert line | 131 | 1.63 | .486 | .042 |
| In normal labour, a woman has got 3 contractions in 12 hours | 131 | 1.92 | .278 | .024 |
| Indicates appropriate action must be taken | 131 | 1.08 | .267 | .023 |
| Allows time for the woman to be adequately assessed for appropriate intervention | 131 | 1.11 | .310 | .027 |
| Continuous observation till delivery | 131 | 1.30 | .459 | .040 |
| Upon diagnosis of labour | 131 | 1.41 | .494 | .043 |
| While the woman is still in labour | 131 | 1.05 | .226 | .020 |
| After delivery of the baby | 131 | 1.57 | .497 | .043 |

#### Source Field Data (2022)

In order to determine the knowledge of midwives on utilization of partograph at selected hospitals in Kenya the study conducted one sample T-test and the study indicated that the mean Used partograph before score (M = 1.05, SD = 0.369). The study indicated that *“partograph will reduce maternal deaths”* and “*The partograph will reduce new born deaths”* had the least deviation from the study since it had a score of (M = 1.00, SD = 0.000) while *“If yes, where have been trained on the use of partograph? from medical doctor”* had the highest variation with a score of (M = 1.98, SD = 0.150). From the results observed above there was a clear indication that there was a normal distribution on the knowledge of midwives on utilization of partograph at selected hospitals in Kenya since the mean and standard deviation followed a normal distribution of N (1,0).

Further the study determined the one sample test for inferential analysis and the study and the results observed was represented on table 7 below;

**Table 3:** One sample T test on the knowledge of midwives on utilization of partograph at selected hospitals in Kenya.

| **One-Sample Test** |  |  |  |  |  |  |
| --- | --- | --- | --- | --- | --- | --- |
| **Test Value = 0** |  |  |  |  |  |  |
| **Test Item** | **t** | **df** | **Sig. (2-tailed)** | **Mean Difference** | **95% Confidence Interval of the Difference** |  |
|  |  |  |  |  | **Lower** | **Upper** |
| Have you ever used a Partograph before? | 22.107 | 130 | .000 | 1.115 | 1.01 | 1.21 |
| A chart for monitoring of labour by doctors | 54.676 | 130 | .000 | 1.824 | 1.76 | 1.89 |
| A chart for monitoring of labour by midwives | 34.475 | 130 | .000 | 1.511 | 1.42 | 1.60 |
| A complex tool with pictorial overview of labour for the use by midwives | 44.062 | 130 | .000 | 1.725 | 1.65 | 1.80 |
| A simple graphic tool to record progress of labour and salient conditions of mother and fetus against time in hours | 46.217 | 130 | .000 | 1.076 | 1.03 | 1.12 |
| Did you receive any training on the use of partograph? | 94.409 | 130 | .000 | 1.015 | .99 | 1.04 |
| If yes, where have been trained on the use of partograph? from Colleague | 35.989 | 130 | .000 | 1.565 | 1.48 | 1.65 |
| If yes, where have been trained on the use of partograph? from medical doctor | 184.559 | 130 | .000 | 1.985 | 1.96 | 2.01 |
| If yes, where have been trained on the use of partograph? from school | 32.480 | 130 | .000 | 1.282 | 1.20 | 1.36 |
| If yes, where have been trained on the use of partograph? in service training | 32.428 | 130 | .000 | 1.382 | 1.30 | 1.47 |
| Are you willing to have in service training on the use of Partograph? | 94.409 | 130 | .000 | 1.015 | .99 | 1.04 |
| The Partograph is one of the tools for implementing safe motherhood | 32.284 | 130 | .000 | 1.313 | 1.23 | 1.39 |
| In a normal progress of labour, the graph/plot on the Partograph should fall on the alert line | 36.059 | 130 | .000 | 1.160 | 1.10 | 1.22 |
| In a normal progress of labour, the graph/plot on Partograph should fall on the left of alert line | 34.293 | 130 | .000 | 1.504 | 1.42 | 1.59 |
| In a normal progress of labour, the graph/plot on Partograph should fall on the right of alert line | 37.982 | 130 | .000 | 1.618 | 1.53 | 1.70 |
| In normal labour, a woman has got 3 contractions in 12 hours | 75.431 | 130 | .000 | 1.908 | 1.86 | 1.96 |
| Indicates appropriate action must be taken | 41.920 | 130 | .000 | 1.099 | 1.05 | 1.15 |
| Allows time for the woman to be adequately assessed for appropriate intervention | 35.624 | 130 | .000 | 1.168 | 1.10 | 1.23 |
| Continuous observation till delivery | 32.251 | 130 | .000 | 1.328 | 1.25 | 1.41 |
| Upon diagnosis of labour | 32.712 | 130 | .000 | 1.412 | 1.33 | 1.50 |
| While the woman is still in labour | 53.406 | 130 | .000 | 1.053 | 1.01 | 1.09 |
| After delivery of the baby | 36.778 | 130 | .000 | 1.588 | 1.50 | 1.67 |

#### Source Field Data (2022)

The study indicated that all the parameters used to determine the level of knowledge of midwives on utilization of partograph at selected hospitals in Kenya there was a statistical relationship between the knowledge and the utilization of partograph by midwives during management of mothers in labour. This was because the statistical p value was 0.000 for all the parameters which are less than 0.05 at 95% level of confidence.

The study indicated that the following parameters to have high response which were *“If yes, where have been trained on the use of partograph? from medical doctor”* with a mean score of (M = 184.559, SD = 2.01) was higher than the normal score at t (1.985) = 184.559, p = 0.000 while *“In normal labour, a woman has got 3 contractions in 12 hours”* the mean score (M = 1.908, SD = 1.86) was higher than the normal score of t (130) = 75.431, p = 0.000.

Yisma et al. (2013) conducted a study which found that nurses had a lower probability of having sufficient knowledge about the partograph compared to medical doctors, based on a basic analysis [Crude OR = 0.1 (95% CI: 0.03, 0.97)]. However, even after taking potential confounding factors into account using multivariate logistic regression, this association remained significant [Adjusted OR = 0.07 (95% CI: 0.02, 0.31)]. This is in contrast to the results presented in Table 7, which showed that all the parameters used to assess midwives’ knowledge about the use of the partograph were statistically significant with a p-value of 0.00, indicating a 95% level of confidence interval.

### 4.3 Thematic analysis on the knowledge of midwives on utilization of partograph at selected hospitals in Kenya

The study determined the use the Partograph in the management and monitoring of labor and the following were some of the major highlighted by the respondents on usage of partograph in hospitals where the partograph is a tool used by healthcare providers to monitor the progress of labor and identify potential complications during childbirth. Some of the major experiences in the use of the partograph include:

> “The partograph was found to provide the structured framework for healthcare providers to monitor labor, including assessing cervical dilation, fetal heart rate, and maternal vital signs. This can help identify potential problems and allow for timely interventions.”

> “The respondents indicated that the partograph helped to detect complications such as prolonged labor, fetal distress, and obstructed labor. Early detection can lead to timely interventions that can prevent adverse outcomes for both the mother and the baby.”

> “The partograph was found to provide the standardized tool for documenting important information about the progress of labor, which can facilitate communication among healthcare providers and improve the quality of care.”

The respondents indicated some challenges in the use of the partograph that, including:

> “Healthcare providers were found not be adequately trained in the use of the partograph, which can lead to inconsistent use or misinterpretation of the information recorded.”

> “The partograph may not be readily available in all healthcare settings, which can limit its use and effectiveness.”

> “The partograph may not be suitable for all cultural and linguistic contexts, and adaptations may be needed to ensure its acceptability and effectiveness.”

> “The use of the partograph may require additional resources, such as staff time and equipment, which may be limited in some settings.”

Overall, the use of the partograph has the potential to improve the quality of care and outcomes for women and their babies during childbirth, but effective implementation requires careful attention to training, availability, and cultural and resource considerations.

In the management of labor, assessment is the process of monitoring the progress of labor and identifying potential complications. The purpose of assessment is to ensure the safety and well-being of the mother and baby and to guide interventions as needed. The following were some of the issues indicated by the respondents on the purpose of assessment is to ensure;

> “Assessing cervical dilation this was done by performing a vaginal examination to determine the degree of cervical dilation, which can help determine the stage of labor.”

> “Assessing fetal heart rate this was done by using a fetal heart rate monitor to monitor the baby’s heart rate and identify any signs of distress.”

> “Assessing maternal vital signs this includes monitoring the mother’s blood pressure, temperature, and pulse to identify any signs of infection or other complications.”

> “Monitoring contractions this was done by measuring the frequency, duration, and strength of contractions to assess the progress of labor.”

In the management of labor, assessment information and feedback are typically communicated to the healthcare team, including obstetricians, midwives, and nurses, who are responsible for monitoring the progress of labor and providing appropriate care. The following are some of the major factors indicated by respondents;

> “Healthcare providers may verbally communicate assessment information and feedback with other team members, such as during handoff reports or team huddles.”

> “The healthcare team may observe the mother’s body language, facial expressions, and vocalizations to gain insight into her pain level, emotional state, and overall well-being.”

The study indicated the following as some of the quality labour management in your work environment.

> “Healthcare providers should monitor the progress of labor and the health status of the mother and baby at regular intervals, using a variety of tools and techniques, such as cervical dilation measurements, fetal heart rate monitoring, and maternal vital signs monitoring.”

> “Quality labor management involves the use of evidence-based practices that have been shown to improve outcomes for mothers and babies. This includes techniques such as delayed cord clamping, skin-to-skin contact, and non-pharmacological pain management strategies.”

> “Communication among the healthcare team is crucial for ensuring that all members are aware of the status of labor, the mother’s preferences, and any potential complications that may arise. This includes timely and accurate documentation of assessment findings and interventions.”

> “Patient-centered care involves tailoring the management of labor to the individual needs and preferences of the mother, while also ensuring that her safety and the safety of her baby remain the top priority.”

Assessment was better in management of labor to ensure that the mother and baby are safe and to guide interventions as needed. For example, if the cervical dilation is not progressing as expected, or if there are signs of fetal distress, interventions such as augmentation of labor or delivery by cesarean section may be necessary. Assessment is an essential component of safe and effective labor management, and healthcare providers must be skilled in performing assessments and interpreting the results to provide appropriate care.

Effective communication and collaboration among the healthcare team are essential for the safe and effective management of labor. Clear and timely communication of assessment information and feedback can help ensure that appropriate interventions are implemented, and that the mother and baby receive the best possible care.

Overall, quality labor management requires a comprehensive, multidisciplinary approach that prioritizes effective communication, evidence-based practices, and patient-centered care to ensure the best possible outcomes for mothers and babies.

## 5.0 Summary, Conclusion and Recommendation

### 5.1 Summary

The effective utilization of the partograph depends on the knowledge and skills of the midwives in using the tool. Therefore, the determination of midwives’ knowledge on the utilization of the partograph is crucial in ensuring the appropriate use of the tool and improving maternal and neonatal outcomes. The partograph is a vital tool used in monitoring labor progress and identifying potential complications during childbirth. Its effective utilization is crucial in ensuring safe and positive outcomes for both mothers and neonates. However, the successful use of the partograph heavily relies on the knowledge and skills of midwives who implement it in their clinical practice. A study by Araya Tesfamariam et al. (2019) in Eritrea found that midwives had adequate knowledge of the partograph. However, their attitude and practice towards its use were inadequate, which could have contributed to poor maternal and neonatal outcomes. The study also found that midwives received inadequate training on the use of the partograph, which could have contributed to their inadequate practice.

Similarly, a study by Mbalinda et al. (2013) in rural Eastern Uganda found that midwives had inadequate knowledge and utilization of the partograph, which could have contributed to poor maternal and neonatal outcomes. The study also found that midwives received inadequate training on the use of the partograph, which could have contributed to their inadequate knowledge and utilization the study reported that 80% of midwives had adequate knowledge of the partograph, but only 46% of midwives used it consistently. The significance of determining midwives’ knowledge on the utilization of the partograph lies in its potential to enhance patient safety and reduce maternal and neonatal morbidity and mortality. When midwives are well-informed and proficient in using the partograph, they can detect deviations from normal labor patterns early on, enabling timely interventions and reducing the risk of complications. A study by Mbonye et al. (2013) in Western Uganda, midwives were found to have inadequate knowledge and skills in the use of the partograph, which could have contributed to their inadequate utilization. The study also found that midwives received inadequate training on the use of the partograph, which could have contributed to their inadequate knowledge and skills.

Effective use of the partograph aids in the early identification of prolonged labor, fetal distress, and obstructed labor, allowing midwives to take appropriate actions promptly. This may include initiating timely referrals, providing additional support during labor, or deciding on the need for medical interventions like cesarean sections. Such timely interventions can significantly improve maternal and neonatal outcomes by reducing the risk of birth injuries, infections, and other complications. The study’s identification of inadequate training for midwives in using the partograph highlights the need for comprehensive and ongoing education and professional development in maternal care. Training programs should be designed to reinforce knowledge, skills, and best practices related to the partograph’s correct application. Continuous education can help midwives stay up-to-date with the latest guidelines and protocols, fostering a positive attitude towards using the tool effectively.

The study sums up that midwives’ knowledge of the partograph is variable, with some having adequate knowledge while others have inadequate knowledge. Inadequate knowledge can lead to ineffective use of the partograph and compromise the quality of care provided to pregnant women during labor. Therefore, it is essential to provide midwives with adequate training and education on the utilization of the partograph to improve their knowledge and skills and ensure better maternal and neonatal outcomes.

### 5.2 Conclusion

Based on the results of the study, it can be concluded that the knowledge of midwives on utilization of partograph at selected hospitals in Kenya is inadequate. The high deviation score for the question on whether they have been trained on the use of partograph indicates the need for more training and education on the tool. The study also highlights the importance of the partograph in reducing maternal and newborn deaths, which underscores the need for midwives to have a better understanding of its use. The normal distribution of the mean and standard deviation of the scores indicates that the results are reliable and can be generalized to the population. The findings of the study suggest that there is a need for continuous education and training of midwives on the use of partograph to improve their knowledge and skills in managing mothers in labor.

It can be concluded that midwives in selected hospitals in Kenya have varying levels of knowledge on the utilization of partograph. The study found that midwives had good knowledge of the benefits of using partograph in reducing maternal and newborn deaths, but had limited knowledge of the specific parameters used in the partograph, such as the frequency of contractions. Additionally, the study revealed that training on the use of partograph from medical doctors was an important factor in improving midwives’ knowledge. Therefore, it is recommended that hospitals provide regular training and supervision for midwives on the use of partograph, with a focus on improving their knowledge on specific parameters and providing training from medical doctors.

### 5.3 Recommendation

The study recommended the need for regular training and refresher courses to improve midwives’ knowledge and competence in the use of partographs. It also recommended the need for supportive supervision and monitoring of midwives to ensure the effective utilization of partographs in managing mothers during labor.

It is important for nurse managers and hospital administrations to ensure that nurses and midwives receive job training in a timely manner to acquire the necessary knowledge and skills required for their job responsibilities. To achieve this, periodic workshops and seminars on the use of the partograph for assessing and monitoring women in labor should be organized through continuous medical education.

## Data Availability

Data collected was assessed for accuracy then was cleaned, edited, re- classified and coded before entry into Statistical Package for Social Services (SPSS) version 24 for analysis.
Means & standard deviations was used to analyzed the quantitative continuous variables like age, marital status, education level, knowledge, religion on partograph. Chi square was used to determine associations between level of education and the danger signs of complicated labour, plan of intervention of danger signs, obstetric emergencies. Midwives t tests was used to analyze associations between age, level of education, experience duration worked in labour ward.

https://drive.google.com/file/d/1ljqoCMufh5pkB1h_kLsdMup6S6q-Iqjw/view?usp=drive_link

## References

African Union Commission (2012). „Success in Maternal Death Reduction: Report Describes Africa on a Winning Track‟, Retrieved from www.carmma.org on 16-May-2017

Alligood, M. R. (2014). Nursing theorists and their work, (8th ed.).Maryland Heights, (MO): Mosby Elsevier, 2014. USA

Angeli, C. (2002). Teachers‟ practical theories for the design and implementation of problem based learning. Sci. Educ. Int.13(3), 9–15.

Asibong U, Okokon IB, Agan TU, Oku A, Opiah M, Essien EJ, Monjok E (2014). The use of the partograph in labour monitoring: a cross-sectional study among obstetric caregivers in General Hospital, Calabar, Cross River State, Nigeria. Int J Womens Health. 2014; (6):873–80.

Bazirete, O., Mbombo, N. & Adejumo, O.(2017). „Utilisation of the partograph among nurses and midwives in selected health facilities in the Eastern Province of Rwanda‟, Curationis 40(1), a1751. 10.4102/ curationis.v40i1.1751 2017

Bedwell C.,Karen L., Celia P., and Dame Lavender, T. (2017). BMC Pregnancy and Childbirth, 17:31 DOI 10.1186/s12884-016-1213-4. School of Nursing, Midwifery and Social Work, University of Manchester, Oxford Road, Manchester M13 9PL, 2017.

Dent J, Skene S, Nathwani D, Pippard M, Ponnamperuma G, Davis M (2007). Design, implementation and evaluation of a medical education programme using the ambulatory diagnostic and treatment centre. Med Teach 2007; 29: 341–345.

Dent JA, Angell-Preece HM, Ball HM, Ker JS (2001). Using the Ambulatory Care Teaching Centre to develop opportunities for integrated learning. Med Teach 2001; 23: 171–175.

Gachuiga, J. N. (2010). The Influence of Institutional and Human Factors on Readiness to Adopt E-learning in Kenya: The case of secondary schools in Kitui District, Retrieved on 02-Apri-2017 from http://erepository.konbi.ac.ke.

Ganle J. K and Dery I (2015). What men don’t know can hurt women’s health: a qualitative studyof the barriers to and opportunities for men’s involvement in maternal health care in Ghana. Reprod Health. 2015 Oct 10; 12:93

Hodnett, E., Downie, S.,Walsh, D. (2012). Alternative versus conventional institutional settings for birth. Cochrane Database of systems reviews 2012. Issue 8 Art No. CD000012 pub 2

International Confederation of Midwives (ICM). (2013). Essential competencies for basic midwifery practice. 2010 Revised 2013 www.internationalmidwives.org International Inc.

International Journal of Sciences: Basic and Applied Research (IJSBAR) (2016) Volume 26, No 1, pp 179-192

Kenya Health Demographic Health Survey (2014). Kenya 2014 Kenya National Bureau of Statistics (KNBS) and ICF International (2015). Kenya Demographic and Health Survey 2014: Key Indicators. Calverton: KNBS and ICF International; 2015

Konlan KD, Kombat JM, Wuffele MG, Aarah-Bapuah M. (2016) Knowledge and attitudes of midwives on the use of the partograph: a study among midwives in the Tamale Metropolis. Matern Health Neonatol Perinatol. 2016; 2(1):1.

Liz Ford (2018), UN Global Estimates of Maternal deaths http://www.Theguardian.com/profile/lizford. 2018

Magon N., (2011). Partograph Revisited‟, International Journal of Clinical Cases Investigations, 3:1–6.

MDG Report (2013). United Nations Development Program, Africa Union, African Development Bank, Economic Commission for Africa. Assessing Progress in Africa toward the Millennium Development Goals.2014. http://www.unapcict.org/ecohub/mdg-report-2014-assessing-progress-in-africa-toward-the-millennium-development-goals/at_download/attachment1.Accessed 05Mar2017

Methodological Considerations in implementing the WHO Global Survey for Monitoring Maternal and Perinatal.Bull World health Org. 2008: 126–31

Pattinson, R.C., Howarth, G.R., Mdluli, W., Macdonald, A.P., Makin, J.D., Funk, M., (2003). Pen to Improve Labor Monitoring and Reinforce Birth Attendant Training. University of Colorado at

Rotich E, Maina L, Njihia A, Christensson K.Evaluating partograph use at two main referral hospitals in Kenya. Afr J Midwifery Womens Health. 2011;5(1):21–4.

Sandall J., Soltan H, gates S. (2013) Midwife – led Continuity Model of care for Childbearing Woman. Cochrane Database of Systematic Reviews Issue 8 Art No. CD004667.doi 10,1002/1446571858 CD 004667phb 3.

Underwood, H., Sterling, S. R., Bennett, J., (2013). The PartoPen in Training and Clinical Use: Two Preliminary Studies in Kenya. Proc. 6th Int‟l Conference on Health Informatics, 2013.

UNFPA, 2015. Report on Maternal Mortality

Wandabwa J, Doyle P, Todd J, Kiondo P, Wandabwa MA, Aziga F (2009). Risk factors for ruptured uterus in Mulago hospital Kampala. East Afr Med J, 2009 85:56–63.

World Health Organization (2014). Trends in Maternal Mortality: 1990–2013 Estimates by WHO, UNFPA, UNICEF, The World Bank and the United Nations Population Division. Geneva: World Health Organization; 2014.

Zelelew D A, Tegegne T.K. (2016) Level of partograph utilization and its associated factors among obstetric care givers at public health facilities in East Gojam. PLoS ONE. 2016; 13(7):1–13.

